# Estimated cardiorespiratory fitness and incident cardiometabolic multimorbidity: findings from three nationally representative cohorts

**DOI:** 10.64898/2026.09.10.26362802

**Authors:** Huan Feng, Tianyue Zhao, Zhengwei Xie, Shanshan Li

## Abstract

**Objective:** Cardiorespiratory fitness is a modifiable marker of cardiometabolic health, but its association with cardiometabolic multimorbidity (CMM) and the generalisability of that association across populations remain unclear. We examined the association between estimated cardiorespiratory fitness (eCRF) and incident CMM in three nationally representative cohorts.

**Materials and methods:** We used data from the China Health and Retirement Longitudinal Study, the English Longitudinal Study of Ageing, and the US Health and Retirement Study. eCRF was derived from a validated sex-specific non-exercise equation and modelled per 1-SD increment and as sex-specific categories. CMM was defined as the coexistence of two or more of hypertension, diabetes, heart disease, and stroke. Associations were assessed using Cox proportional hazards models, with supplementary analyses of proportional hazards, competing death, multiple imputation, prediction performance, and meta-analysis.

**Results:** Among 12,944 participants, 3,029 developed incident CMM. Each 1-SD increment in eCRF was associated with a lower risk in the pooled model (hazard ratio 0.61, 95% CI 0.58-0.64). Compared with low fitness, high fitness was associated with a substantially lower risk (hazard ratio 0.40, 95% CI 0.35-0.45; P for trend <0.001). Adding eCRF to sociodemographic and behavioural predictors improved discrimination, and the association remained directionally consistent across sensitivity analyses.

**Conclusions:** Higher estimated cardiorespiratory fitness was independently associated with a lower risk of incident CMM across three diverse populations and provided modest incremental prognostic information. As an easily obtained, non-exercise measure, eCRF may help characterise cardiometabolic risk in ageing populations, although between-cohort heterogeneity and the observational design warrant cautious interpretation.

## 1 Introduction

Global population ageing has produced a steep rise in multimorbidity, and more than half of adults aged 60 years and older now live with two or more chronic conditions ^[1]^. Cardiometabolic multimorbidity, the coexistence of two or more cardiometabolic disorders such as hypertension, diabetes, heart disease, and stroke, is among the most harmful patterns ^[2]^. Its components share common antecedents and tend to cluster, and when they co-occur the risks multiply: in pooled data from nearly 700,000 participants, a history of any two of diabetes, stroke, and myocardial infarction at age 60 years was associated with roughly 12 years of reduced life expectancy ^[3]^. Cardiometabolic multimorbidity also accelerates cognitive decline and disability ^[4]^, and is expected to grow fastest in low- and middle-income countries undergoing rapid lifestyle transition ^[5]^. Accessible, modifiable markers that identify high-risk individuals before multimorbidity becomes established are therefore needed.

Cardiorespiratory fitness is one such candidate. It is among the strongest predictors of cardiovascular disease and mortality, often outperforming traditional risk factors ^[6]^, and the relationship is graded: each one-metabolic-equivalent (MET) increment is associated with an approximately 13% lower risk of all-cause mortality and coronary events ^[7]^, a pattern confirmed in quantitative meta-analysis ^[8]^. Fitness is also inversely and dose-dependently related to incident type 2 diabetes independently of adiposity ^[9]^, and Mendelian randomisation evidence indicates that this association is at least partly causal ^[10]^. These effects are biologically coherent, since higher fitness is accompanied by greater insulin sensitivity, more favourable lipid and glucose profiles, lower visceral adiposity, and improved vascular and autonomic function ^[6]^. Because these pathways act on the shared roots of cardiometabolic disease, fitness is well placed to influence not only individual conditions but their co-occurrence; consistent with this, higher directly measured fitness predicted slower progression from health to a first cardiometabolic disease and onward to multimorbidity in the UK Biobank ^[11]^. A major obstacle is that fitness is seldom measured. The reference standard, maximal oxygen uptake from graded exercise testing, requires specialised equipment and effort and is often impractical for older adults, leaving fitness the only major risk factor not routinely recorded in primary care; accordingly, the American Heart Association has recommended that fitness be treated as a clinical vital sign and, where exercise testing is unavailable, estimated from non-exercise prediction equations ^[12]^. Such equations derive fitness from age, sex, body mass index, waist circumference, resting heart rate, physical activity, and smoking, and estimated cardiorespiratory fitness (eCRF) predicts cardiovascular events and mortality comparably to measured fitness ^[13]^. eCRF has been linked to incident hypertension in initially normotensive adults ^[14]^ and, in cross-national work, to hypertension and mortality across American and Chinese populations ^[15]^, to arthritis across four cohorts ^[16]^, and to depressive symptoms in the United States, England, and China ^[17]^, although these studies addressed single conditions and reported associations that sometimes differed by country.

To the best of our knowledge, however, no study has examined whether eCRF predicts the onset of cardiometabolic multimorbidity, and whether such an association is generalisable across diverse populations remains unclear. Considering that cardiometabolic multimorbidity has become an increasingly important public health priority in ageing populations, clarifying the association between eCRF and cardiometabolic multimorbidity could offer valuable insights for its early identification and prevention. Several nationwide, population-based, prospective cohorts have collected data on older populations, including the China Health and Retirement Longitudinal Study (CHARLS), the English Longitudinal Study of Ageing (ELSA), and the Health and Retirement Study (HRS). In this study, by using nationally representative samples of older adults, we aim to explore the relationship between eCRF and the incidence of cardiometabolic multimorbidity. These findings may help refine risk stratification and inform targeted prevention strategies for cardiometabolic multimorbidity in ageing populations.

## 2 Methods

### 2.1 Study design and study population

This study was a harmonised analysis of three nationally representative, prospective cohorts of community-dwelling middle-aged and older adults: CHARLS ^[18]^, ELSA ^[19]^, and HRS ^[20]^. ELSA and CHARLS were established as sister studies of HRS and share its design, sampling framework, and core instruments, ensuring strong cross-cohort comparability. CHARLS used 2011 as the baseline, with follow-up through 2018; ELSA used Wave 2 (2004/05), with follow-up through Wave 8 (2016/17); and HRS used Wave 8 (2006), the first wave collecting the physical measurements needed to estimate fitness, with follow-up through Wave 15 (2020). The minimum age at entry was 45 years for CHARLS and 50 years for ELSA and HRS. Each cohort obtained ethical approval and written informed consent in its respective country; because only de-identified and openly accessible data were used, no further ethical review was sought for the present analysis.

For each cohort, we applied an identical selection procedure (Figure 1). Beginning with all baseline respondents, we excluded those below the cohort-specific minimum age, those with missing or implausible values for the variables required to estimate cardiorespiratory fitness, and those with missing covariate data, yielding the cross-sectional analytical sample for the fully adjusted analysis (n = 16,696; CHARLS, 4,860; ELSA, 5,394; HRS, 6,442). For the longitudinal analysis, we further excluded participants with cardiometabolic multimorbidity at baseline and those without any follow-up assessment, leaving 12,944 participants in the fixed fully adjusted complete-case risk set (CHARLS, 4,208; ELSA, 4,257; HRS, 4,479).

**Figure 1.**
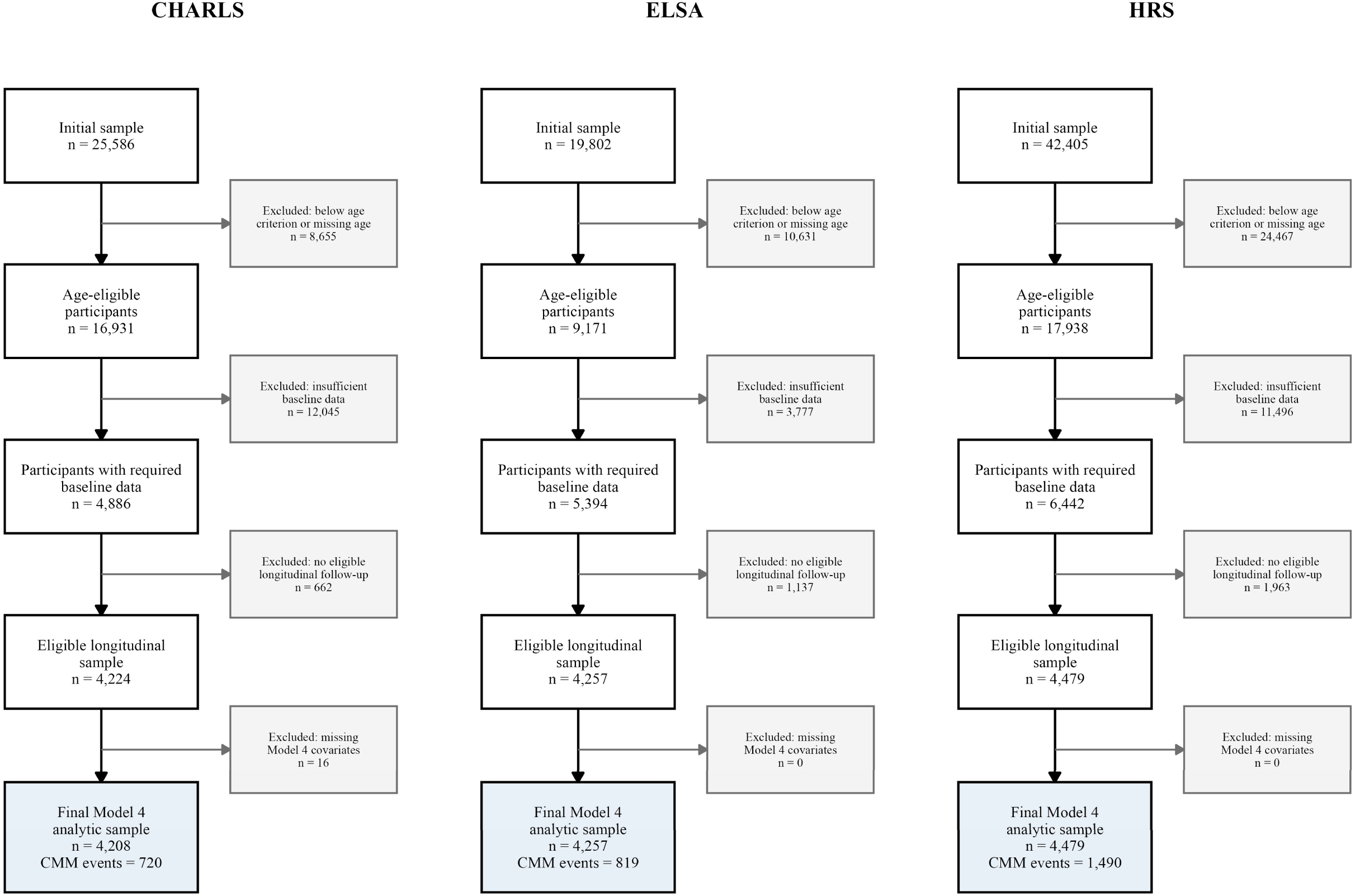
Flow chart of participant selection in CHARLS, ELSA, and HRS.

### 2.2 Estimated cardiorespiratory fitness

We derived CRF from a previously validated, sex-specific equation that does not require exercise testing; this equation converts age, BMI, waist circumference, resting heart rate, physical activity, and smoking status into a value expressed in METs ^[12]^. This approach is well supported by recent large-scale evidence: in a meta-analysis of 42 studies comprising 35 cohorts and 3.8 million observations, the predictive strength of equation-based CRF for death from any cause and from cardiovascular causes was comparable to that of CRF obtained through exercise testing, indicating that eCRF is a practical and robust alternative for risk stratification across diverse populations ^[21]^. The same equation has also forecast mortality outcomes in population-representative surveys ^[22]^.

The equations were specified as follows ^[12]^: for men, eCRF (METs) = 21.2870 + 0.1654 × age − 0.0023 × age² − 0.2318 × BMI − 0.0337 × waist circumference − 0.0390 × resting heart rate + 0.6351 × physically active − 0.4263 × current smoker; for women, eCRF (METs) = 14.7873 + 0.1159 × age − 0.0017 × age² − 0.1534 × BMI − 0.0085 × waist circumference − 0.0364 × resting heart rate + 0.5987 × physically active − 0.2994 × current smoker. Age, BMI, waist circumference (cm), and resting heart rate (beats per minute) were entered as continuous variables, whereas physical activity and current smoking were entered as dichotomous variables (1 if present, 0 if absent). The same equation and harmonisation rules were applied uniformly across the three cohorts, and estimated values falling outside a plausible physiological range (4–20 METs) were treated as missing. Three parameterisations of eCRF were used: a standardised continuous form (mean 0, SD 1; reported per 1-SD increase); a categorical form based on sex-specific quintiles and collapsed into low (quintile 1, reference), moderate (quintiles 2–3), and high (quintiles 4–5) fitness; and, for the dose–response analysis, as a continuous variable modelled with restricted cubic splines.

### 2.3 Cardiometabolic multimorbidity

Following commonly applied criteria, we considered CMM present when a participant had at least two of the following four conditions: hypertension, diabetes, heart disease, or stroke in previous cohort studies ^[3, 11]^. Each condition was ascertained from self-reported physician diagnosis using harmonised items available in all three cohorts. Participants were classified as having CMM when at least two of the four conditions were present, as free of CMM when fewer than two were present, and as having missing status only when information on all four conditions was unavailable.

For the longitudinal analysis, only participants free of CMM at baseline were included, and incident CMM was identified as the first follow-up wave at which a participant first satisfied the criterion of two or more coexisting conditions. Follow-up time was assigned according to the wave intervals specific to each cohort, with the event time corresponding to the wave of first diagnosis and censoring at the last available assessment for those who remained free of CMM.

### 2.4 Covariates

The covariates included in this analysis were educational attainment (lower secondary or below versus upper secondary or above), labour-force status (working, retired, or unemployed/out of the labour force), marital status (married or partnered versus other), living alone (no versus yes), smoking (never, former, or current), alcohol consumption (non-drinker versus drinker, based on whether the participant had consumed alcohol in the past year), restless sleep (no versus yes), disability in activities of daily living (ADL) and instrumental activities of daily living (IADL), and physician-diagnosed comorbidities. Functional status was assessed using six-item ADL and five-item IADL scales that were broadly comparable but not identical across cohorts [23]; for both, participants reporting any difficulty were classified as having disability and those reporting none as having no disability, and ADL and IADL disability were adjusted for in all three cohorts. Comorbidities comprised arthritis, cancer, and lung disease, with kidney and liver disease additionally adjusted for in CHARLS; area of residence (urban versus rural) was additionally available in CHARLS. Age was modelled as a continuous variable in all regression analyses, whereas the categorical age grouping (50–64 and ≥65 years; 45–64 and ≥65 years in CHARLS) was used only for the age-stratified subgroup analysis. A detailed description of all covariates is provided in Supplementary Table S3.

### 2.5 Statistical analysis

Baseline characteristics were summarised as means with standard deviations (SD) for continuous variables and as frequencies with percentages for categorical variables, and were compared across cohorts using one-way analysis of variance and chi-squared tests, as appropriate. eCRF was analysed both as a continuous variable (per 1-SD increment) and as sex-specific categories (low, moderate, and high).

Four sequentially adjusted models with an identical structure were fitted in each cohort: Model 1 was unadjusted; Model 2 was adjusted for age and sex; Model 3 was additionally adjusted for educational attainment, labour-force status, marital status, living alone, smoking, alcohol consumption, and restless sleep; and Model 4 was additionally adjusted for ADL and IADL disability, arthritis, cancer, and lung disease (and, in CHARLS, kidney and liver disease and area of residence). Cross-sectional associations between eCRF and prevalent CMM were assessed using logistic regression and reported as odds ratios (ORs) with 95% confidence intervals (CIs). Prospective associations between baseline eCRF and incident CMM were assessed using Cox proportional hazards regression, with follow-up time as the time scale, and reported as hazard ratios (HRs) with 95% CIs. Pooled Cox models used cohort-stratified baseline hazards. Linear trends across categories were tested by modelling the ordinal category as a continuous term.

Potential non-linear dose-response relationships between eCRF and incident CMM were examined using restricted cubic splines, with the cohort-specific median eCRF as the reference. Effect modification was evaluated in prespecified subgroups and by likelihood-ratio tests for eCRF-by-cohort, eCRF-by-age, and eCRF-by-sex interactions. The proportional-hazards assumption was assessed using scaled Schoenfeld residuals, and models addressing detected departures were fitted when needed. Additional analyses evaluated death as a competing event, multiple imputation for missing covariates, common-covariate adjustment, within-cohort relative eCRF rank, a common 7-year follow-up window, per-MET scaling, leave-one-cohort-out estimates, incremental prediction performance, and two-stage random-effects meta-analysis. Details are provided in the Supplementary Methods. Proportional-hazards diagnostics and the models addressing detected departures are reported in Supplementary Tables S4-S6 and Supplementary Figure S3. All analyses were performed in R, and a two-sided P value <0.05 was considered statistically significant.

## 3 Results

### 3.1 Baseline characteristics

The cross-sectional fully adjusted analytical sample comprised 16,696 participants (4,860 from CHARLS, 5,394 from ELSA, and 6,442 from HRS). After excluding those with CMM at baseline and those without follow-up, 12,944 participants were included in the fixed fully adjusted longitudinal risk set (4,208 from CHARLS, 4,257 from ELSA, and 4,479 from HRS), and the participant selection process is shown in Figure 1. Among the longitudinal sample, the overall mean age was 63.1 (SD 9.7) years, and 7,230 participants (55.9%) were women. Baseline characteristics differed markedly across cohorts (Table 1): CHARLS participants were the youngest, HRS participants the oldest, and mean eCRF was highest in CHARLS, intermediate in ELSA, and lowest in HRS. Missingness, included-versus-excluded comparisons, and variable definitions are shown in Supplementary Tables S1-S3.

**Table 1.** Baseline characteristics of the longitudinal cohorts.

| Characteristic | Category | CHARLS | ELSA | HRS | Pooled |
| --- | --- | --- | --- | --- | --- |
| Age, years | Mean (SD) | 58.30 (9.08) | 64.65 (8.74) | 66.29 (9.49) | 63.15 (9.74) |
| eCRF, METs | Mean (SD) | 10.44 (2.04) | 8.97 (1.98) | 8.34 (1.93) | 9.23 (2.17) |
| Sex | Male | 1907 (45.3%) | 1939 (45.5%) | 1868 (41.7%) | 5714 (44.1%) |
| Sex | Female | 2301 (54.7%) | 2318 (54.5%) | 2611 (58.3%) | 7230 (55.9%) |
| eCRF category | Low (Q1) | 708 (16.8%) | 702 (16.5%) | 678 (15.1%) | 2088 (16.1%) |
| eCRF category | Moderate (Q2-3) | 1683 (40.0%) | 1662 (39.0%) | 1697 (37.9%) | 5042 (39.0%) |
| eCRF category | High (Q4-5) | 1817 (43.2%) | 1893 (44.5%) | 2104 (47.0%) | 5814 (44.9%) |
| Education | Upper secondary or above | 1299 (30.9%) | 2533 (59.5%) | 3771 (84.2%) | 7603 (58.7%) |
| Education | Lower secondary or below | 2909 (69.1%) | 1724 (40.5%) | 708 (15.8%) | 5341 (41.3%) |
| Labour-force status | Working | 2784 (66.2%) | 1519 (35.7%) | 1525 (34.0%) | 5828 (45.0%) |
| Labour-force status | Retired | 1274 (30.3%) | 25 (0.6%) | 2034 (45.4%) | 3333 (25.7%) |
| Labour-force status | Unemployed/Out of LF | 150 (3.6%) | 2713 (63.7%) | 920 (20.5%) | 3783 (29.2%) |
| Marital status | Married/Partnered | 3721 (88.4%) | 3176 (74.6%) | 3036 (67.8%) | 9933 (76.7%) |
| Marital status | Other | 487 (11.6%) | 1081 (25.4%) | 1443 (32.2%) | 3011 (23.3%) |
| Living alone | No | 4020 (95.5%) | 3436 (80.7%) | 3599 (80.4%) | 11055 (85.4%) |
| Living alone | Yes | 188 (4.5%) | 821 (19.3%) | 880 (19.6%) | 1889 (14.6%) |
| Smoking | Never smoker | 2653 (63.0%) | 1641 (38.5%) | 2015 (45.0%) | 6309 (48.7%) |
| Smoking | Former smoker | 321 (7.6%) | 2029 (47.7%) | 1829 (40.8%) | 4179 (32.3%) |
| Smoking | Current smoker | 1234 (29.3%) | 587 (13.8%) | 635 (14.2%) | 2456 (19.0%) |
| Drinking | Non-drinker | 2926 (69.5%) | 342 (8.0%) | 1909 (42.6%) | 5177 (40.0%) |
| Drinking | Drinker | 1282 (30.5%) | 3915 (92.0%) | 2570 (57.4%) | 7767 (60.0%) |
| Restless sleep | No | 2113 (50.2%) | 2513 (59.0%) | 3322 (74.2%) | 7948 (61.4%) |
| Restless sleep | Yes | 2095 (49.8%) | 1744 (41.0%) | 1157 (25.8%) | 4996 (38.6%) |
| ADL disability | No | 3610 (85.8%) | 3663 (86.0%) | 4048 (90.4%) | 11321 (87.5%) |
| ADL disability | Yes | 598 (14.2%) | 594 (14.0%) | 431 (9.6%) | 1623 (12.5%) |
| IADL disability | No | 3402 (80.8%) | 3965 (93.1%) | 3871 (86.4%) | 11238 (86.8%) |
| IADL disability | Yes | 806 (19.2%) | 292 (6.9%) | 608 (13.6%) | 1706 (13.2%) |
| Arthritis | No | 2810 (66.8%) | 2931 (68.9%) | 2222 (49.6%) | 7963 (61.5%) |
| Arthritis | Yes | 1398 (33.2%) | 1326 (31.1%) | 2257 (50.4%) | 4981 (38.5%) |
| Cancer | No | 4173 (99.2%) | 3992 (93.8%) | 3914 (87.4%) | 12079 (93.3%) |
| Cancer | Yes | 35 (0.8%) | 265 (6.2%) | 565 (12.6%) | 865 (6.7%) |
| Lung disease | No | 3805 (90.4%) | 4029 (94.6%) | 4195 (93.7%) | 12029 (92.9%) |
| Lung disease | Yes | 403 (9.6%) | 228 (5.4%) | 284 (6.3%) | 915 (7.1%) |

Notes: Values are mean (SD) or n (%). The fixed Model 4 complete-case samples were used. CMM, cardiometabolic multimorbidity; eCRF, estimated cardiorespiratory fitness; MET, metabolic equivalent; ADL, activities of daily living; IADL, instrumental activities of daily living.

### 3.2 Association between eCRF and CMM

In the fixed fully adjusted longitudinal risk set, 3,029 of 12,944 participants developed incident CMM. Events occurred in 720 of 4,208 CHARLS participants, 819 of 4,257 ELSA participants, and 1,490 of 4,479 HRS participants. Kaplan-Meier curves showed that cumulative CMM incidence was lowest in the high-fitness group and highest in the low-fitness group in every cohort, with clear separation across the three categories (Figure 2).

**Figure 2.**
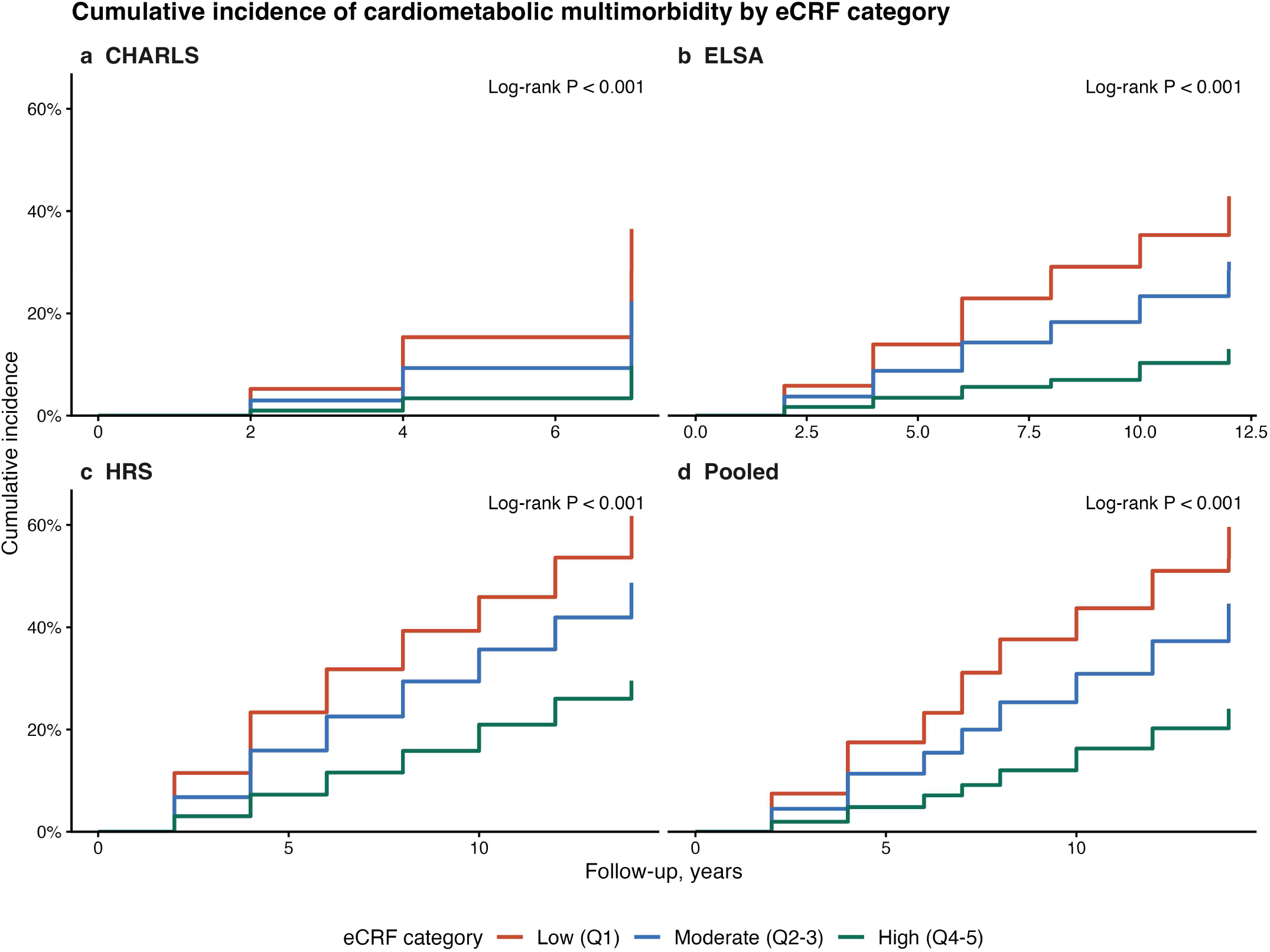
Kaplan–Meier curves for the cumulative incidence of CMM by eCRF category in CHARLS, ELSA, HRS, and the pooled sample. eCRF categories are low (Q1), moderate (Q2–3), and high (Q4–5). P values are from the log-rank test. CMM, cardiometabolic multimorbidity; eCRF, estimated cardiorespiratory fitness; Q, quintile.

Higher eCRF was consistently associated with a lower risk of incident CMM after sequential adjustment (Table 3). In the fully adjusted model, each 1-SD increment in eCRF was associated with an HR of 0.43 (95% CI 0.38-0.49) in CHARLS, 0.63 (95% CI 0.57-0.70) in ELSA, 0.66 (95% CI 0.62-0.72) in HRS, and 0.61 (95% CI 0.58-0.64) in the pooled cohort-stratified model (all P <0.001). When eCRF was modelled as sex-specific categories, participants with high fitness had a lower risk than those with low fitness, with fully adjusted HRs of 0.24 (95% CI 0.19-0.31) in CHARLS, 0.41 (95% CI 0.32-0.52) in ELSA, 0.49 (95% CI 0.42-0.59) in HRS, and 0.40 (95% CI 0.35-0.45) in the pooled sample. Moderate fitness showed intermediate risks, and trends across categories were significant throughout. Cross-sectional analyses were consistent, with a pooled fully adjusted OR of 0.58 (95% CI 0.54-0.62) per 1-SD higher eCRF and 0.32 (95% CI 0.28-0.37) for high versus low fitness (Table 2).

**Table 2.**
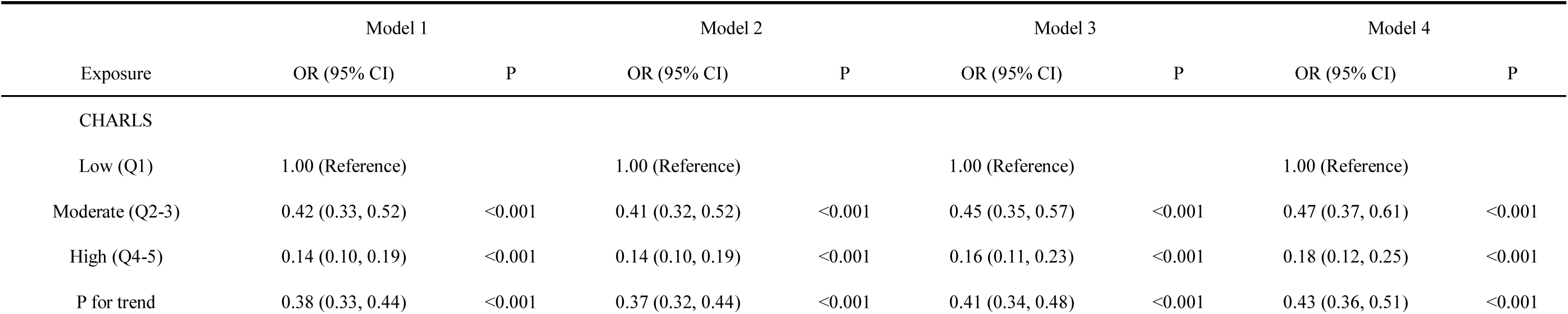
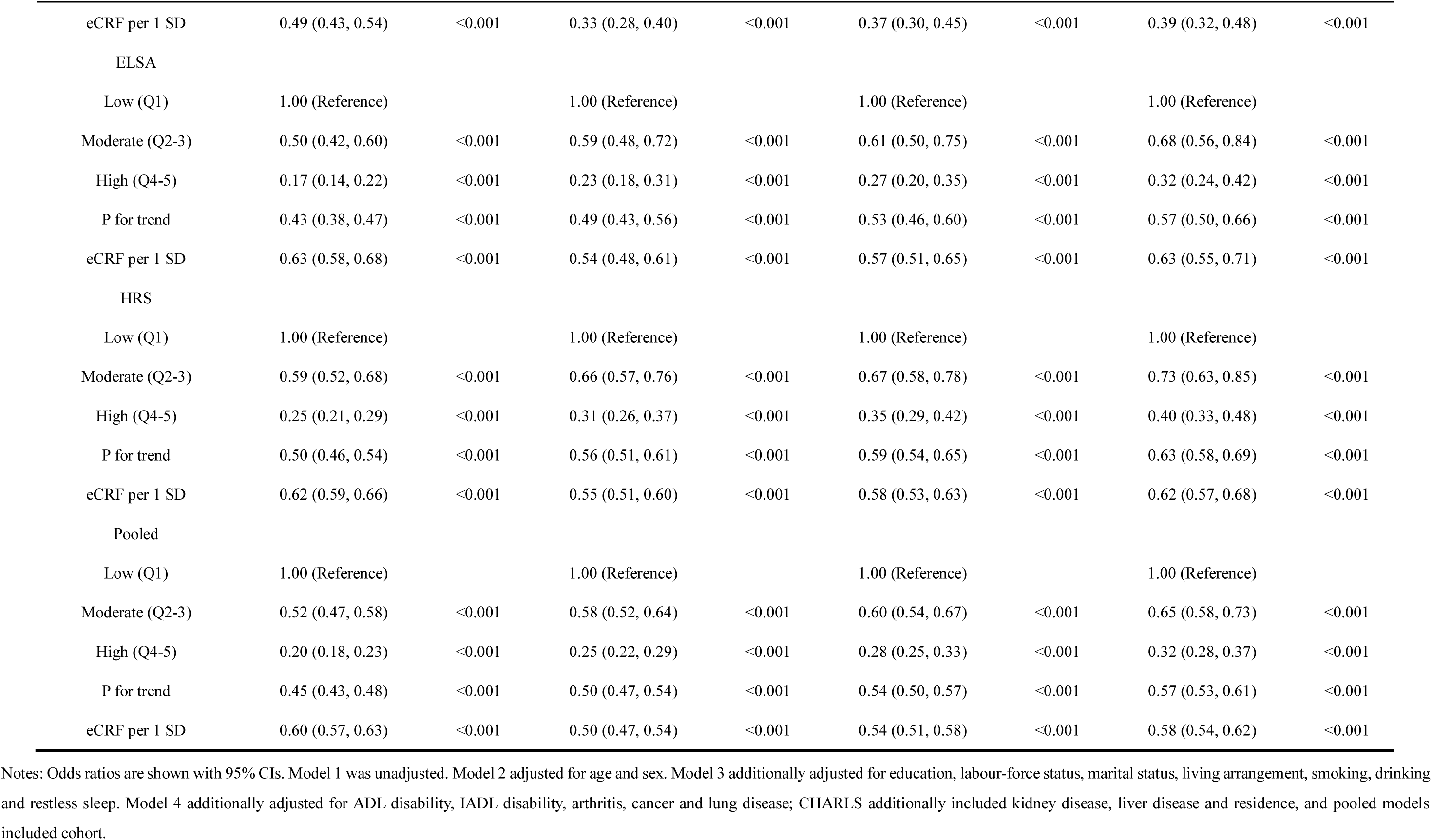
Cross-sectional associations between eCRF and prevalent CMM.

**Table 3.** Prospective associations between eCRF and incident CMM.

| Exposure | Model 1 |  | Model 2 |  | Model 3 |  | Model 4 |  |
| --- | --- | --- | --- | --- | --- | --- | --- | --- |
|  | HR (95% CI) | P | HR (95% CI) | P | HR (95% CI) | P | HR (95% CI) | P |
| CHARLS |  |  |  |  |  |  |  |  |
| Low (Q1) | 1.00 (Reference) |  | 1.00 (Reference) |  | 1.00 (Reference) |  | 1.00 (Reference) |  |
| Moderate (Q2-3) | 0.56 (0.47, 0.66) | <0.001 | 0.57 (0.47, 0.68) | <0.001 | 0.57 (0.47, 0.69) | <0.001 | 0.58 (0.48, 0.70) | <0.001 |
| High (Q4-5) | 0.23 (0.18, 0.28) | <0.001 | 0.23 (0.18, 0.29) | <0.001 | 0.23 (0.18, 0.30) | <0.001 | 0.24 (0.19, 0.31) | <0.001 |
| P for trend | 0.48 (0.44, 0.53) | <0.001 | 0.48 (0.43, 0.54) | <0.001 | 0.49 (0.43, 0.55) | <0.001 | 0.49 (0.43, 0.55) | <0.001 |
| eCRF per 1 SD | 0.64 (0.59, 0.69) | <0.001 | 0.43 (0.38, 0.48) | <0.001 | 0.43 (0.38, 0.49) | <0.001 | 0.43 (0.38, 0.49) | <0.001 |
| ELSA |  |  |  |  |  |  |  |  |
| Low (Q1) | 1.00 (Reference) |  | 1.00 (Reference) |  | 1.00 (Reference) |  | 1.00 (Reference) |  |
| Moderate (Q2-3) | 0.62 (0.52, 0.73) | <0.001 | 0.74 (0.61, 0.88) | 0.001 | 0.76 (0.63, 0.91) | 0.003 | 0.81 (0.67, 0.98) | 0.027 |
| High (Q4-5) | 0.24 (0.20, 0.29) | <0.001 | 0.33 (0.26, 0.42) | <0.001 | 0.37 (0.29, 0.46) | <0.001 | 0.41 (0.32, 0.52) | <0.001 |
| P for trend | 0.49 (0.45, 0.54) | <0.001 | 0.58 (0.51, 0.64) | <0.001 | 0.60 (0.54, 0.68) | <0.001 | 0.63 (0.56, 0.71) | <0.001 |
| eCRF per 1 SD | 0.66 (0.61, 0.71) | <0.001 | 0.57 (0.52, 0.63) | <0.001 | 0.60 (0.54, 0.67) | <0.001 | 0.63 (0.57, 0.70) | <0.001 |
| HRS |  |  |  |  |  |  |  |  |
| Low (Q1) | 1.00 (Reference) |  | 1.00 (Reference) |  | 1.00 (Reference) |  | 1.00 (Reference) |  |
| Moderate (Q2-3) | 0.69 (0.60, 0.79) | <0.001 | 0.77 (0.67, 0.88) | <0.001 | 0.77 (0.67, 0.89) | <0.001 | 0.79 (0.68, 0.91) | 0.001 |
| High (Q4-5) | 0.36 (0.31, 0.41) | <0.001 | 0.46 (0.39, 0.55) | <0.001 | 0.48 (0.40, 0.56) | <0.001 | 0.49 (0.42, 0.59) | <0.001 |
| P for trend | 0.59 (0.55, 0.63) | <0.001 | 0.67 (0.62, 0.73) | <0.001 | 0.68 (0.63, 0.74) | <0.001 | 0.69 (0.64, 0.75) | <0.001 |
| eCRF per 1 SD | 0.70 (0.66, 0.74) | <0.001 | 0.65 (0.60, 0.70) | <0.001 | 0.66 (0.61, 0.71) | <0.001 | 0.66 (0.62, 0.72) | <0.001 |
| Pooled |  |  |  |  |  |  |  |  |
| Low (Q1) | 1.00 (Reference) |  | 1.00 (Reference) |  | 1.00 (Reference) |  | 1.00 (Reference) |  |

|  | Model 1 |  | Model 2 |  | Model 3 |  | Model 4 |  |
| --- | --- | --- | --- | --- | --- | --- | --- | --- |
| Exposure | HR (95% CI) | P | HR (95% CI) | P | HR (95% CI) | P | HR (95% CI) | P |
| Moderate (Q2-3) | 0.63 (0.57, 0.69) | <0.001 | 0.70 (0.64, 0.77) | <0.001 | 0.71 (0.65, 0.78) | <0.001 | 0.74 (0.67, 0.81) | <0.001 |
| High (Q4-5) | 0.29 (0.26, 0.32) | <0.001 | 0.36 (0.32, 0.40) | <0.001 | 0.37 (0.33, 0.42) | <0.001 | 0.40 (0.35, 0.45) | <0.001 |
| P for trend | 0.53 (0.51, 0.56) | <0.001 | 0.60 (0.56, 0.63) | <0.001 | 0.61 (0.57, 0.64) | <0.001 | 0.62 (0.59, 0.66) | <0.001 |
| eCRF per 1 SD | 0.67 (0.65, 0.70) | <0.001 | 0.58 (0.55, 0.61) | <0.001 | 0.59 (0.56, 0.62) | <0.001 | 0.61 (0.58, 0.64) | <0.001 |

Notes: Hazard ratios are shown with 95% CIs. All models used a fixed Model 4 complete-case risk set. The pooled Cox model used cohort-stratified baseline hazards. Covariate adjustment followed the hierarchy defined for Table 2.

Restricted cubic spline analyses demonstrated an inverse dose-response relationship between eCRF and incident CMM across cohorts (Figure 3). The pooled spline suggested lower risk at higher eCRF values, while cohort-specific curves remained directionally consistent. Because non-linearity was more apparent in the pooled sample than within individual cohorts, the spline findings were interpreted as supporting a graded association rather than a single universal threshold; cohort-specific spline sensitivity plots are shown in Supplementary Figure S6.

**Figure 3.**
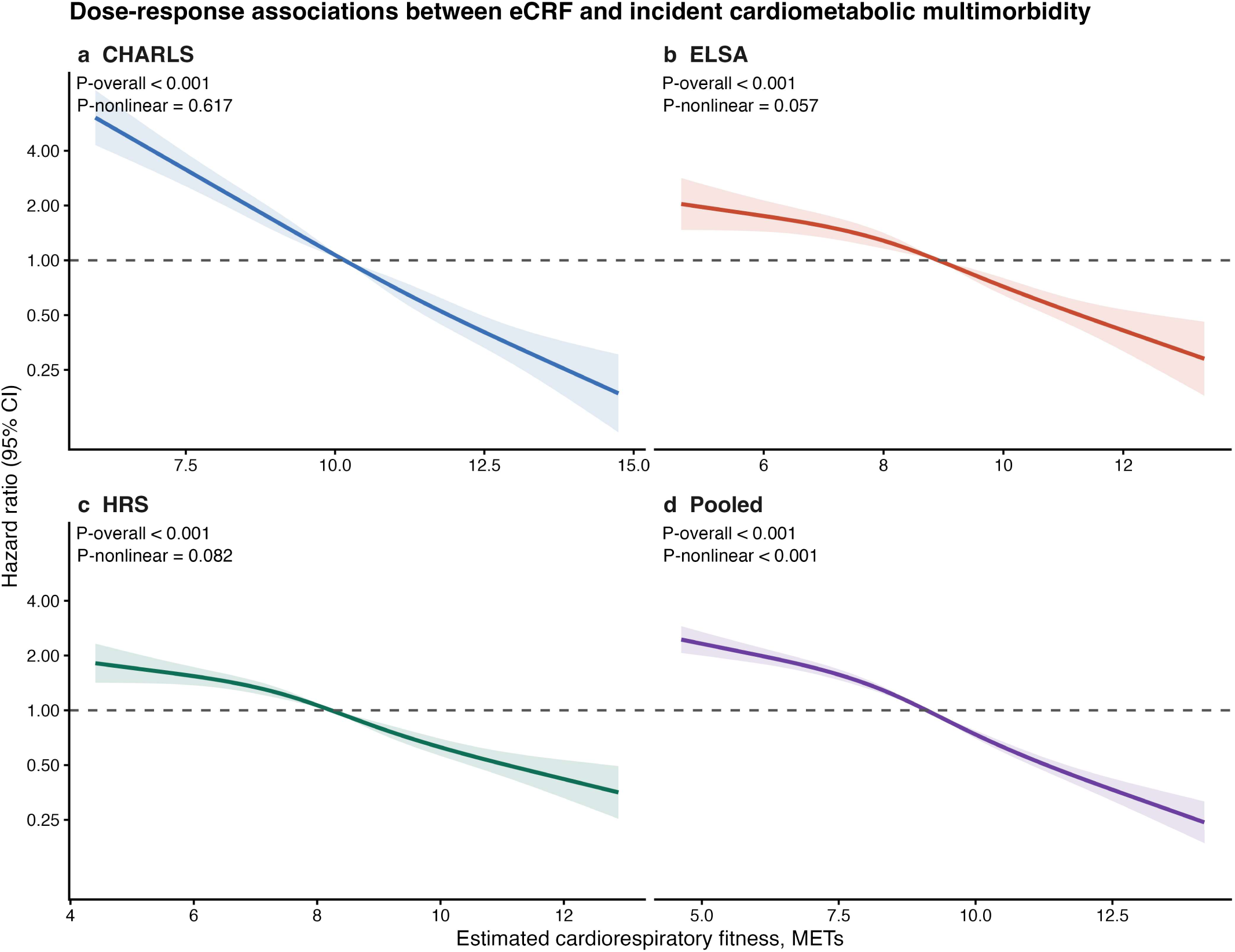
Restricted cubic spline curves for the association between eCRF and incident CMM. Solid lines denote hazard ratios and shaded areas the 95% confidence intervals, with the cohort-specific median eCRF as the reference. Models were fully adjusted. CMM, cardiometabolic multimorbidity; eCRF, estimated cardiorespiratory fitness; MET, metabolic equivalent.

### 3.3 Subgroup analyses and effect modification

Effect modification analyses indicated statistically detectable heterogeneity by cohort, age, and sex. Likelihood-ratio tests gave P = 0.004 for eCRF by cohort, P <0.001 for eCRF by age, and P <0.001 for eCRF by sex (Supplementary Table S12). Stratified estimates remained below unity across key subgroups (Figure 4), but the association was stronger in younger participants and in women. Construct-validity analyses supported the eCRF measure: Spearman correlations between eCRF and grip strength-to-weight ratio were 0.61 (95% CI 0.58-0.63) in CHARLS, 0.74 (95% CI 0.72-0.75) in ELSA, and 0.72 (95% CI 0.70-0.73) in HRS (Supplementary Table S13; Supplementary Figure S4).

**Figure 4.**
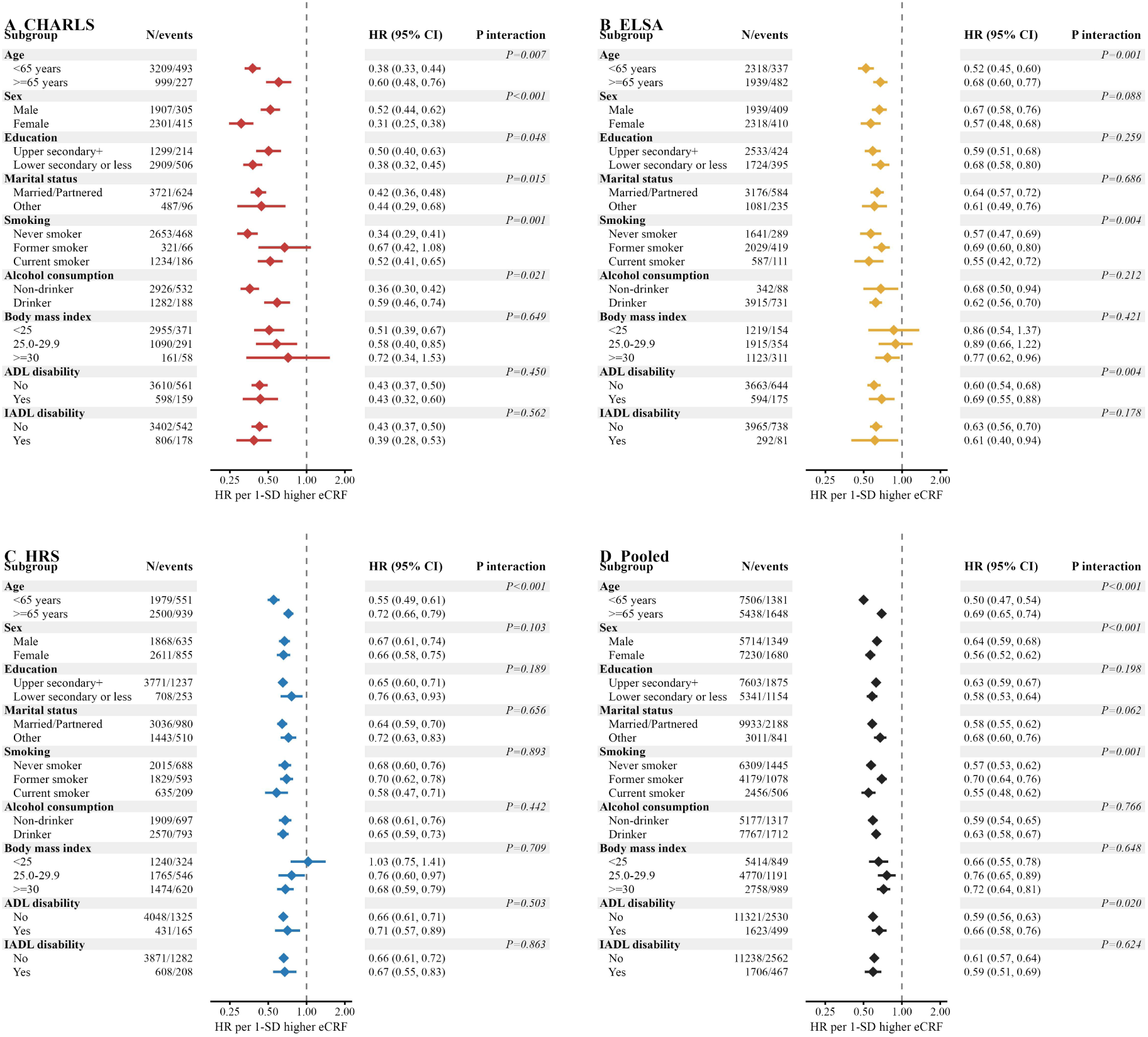
Subgroup analyses and interaction tests for eCRF and incident CMM. Models were fully adjusted. CMM, cardiometabolic multimorbidity; eCRF, estimated cardiorespiratory fitness; HR, hazard ratio; MET, metabolic equivalent.

The main association remained robust across sensitivity analyses. Accounting for death as a competing event gave a pooled subdistribution HR of 0.63 (95% CI 0.60-0.67) per 1-SD higher eCRF, with competing-risk cumulative incidence shown in Supplementary Figure S2. Multiple imputation gave an essentially identical pooled HR of 0.61 (95% CI 0.58-0.64). Restricting all cohorts to a common 7-year follow-up window yielded a pooled HR of 0.57 (95% CI 0.53-0.61), based on 1,966 events; using within-cohort relative rank gave an HR of 0.85 (95% CI 0.84-0.87) per 10-percentile increase; and the per-1-MET estimate was 0.78 (95% CI 0.76-0.80) (Supplementary Tables S7-S14; Supplementary Figure S7). Adding eCRF to Model 3 improved pooled 5-year AUC from 0.658 (95% CI 0.642-0.674) to 0.698 (95% CI 0.683-0.713) and 10-year AUC from 0.707 (95% CI 0.694-0.720) to 0.736 (95% CI 0.724-0.748). The pooled Harrell C-index increment was 0.037 (95% CI 0.030-0.044), and continuous NRI was 0.155 (95% CI 0.122-0.194) at 5 years and 0.204 (95% CI 0.176-0.226) at 10 years (Table 4; Supplementary Table S15; Supplementary Figure S1 and Supplementary Figure S5). Two-stage meta-analysis supported an inverse association but showed substantial heterogeneity: for each 1-SD higher eCRF, REML pooling gave HR 0.57 (95% CI 0.44-0.74), I2 = 95.0%, whereas the Hartung-Knapp interval was wider (HR 0.57, 95% CI 0.32-1.02; Figure 5; Table 5; Supplementary Table S16).

**Figure 5.**
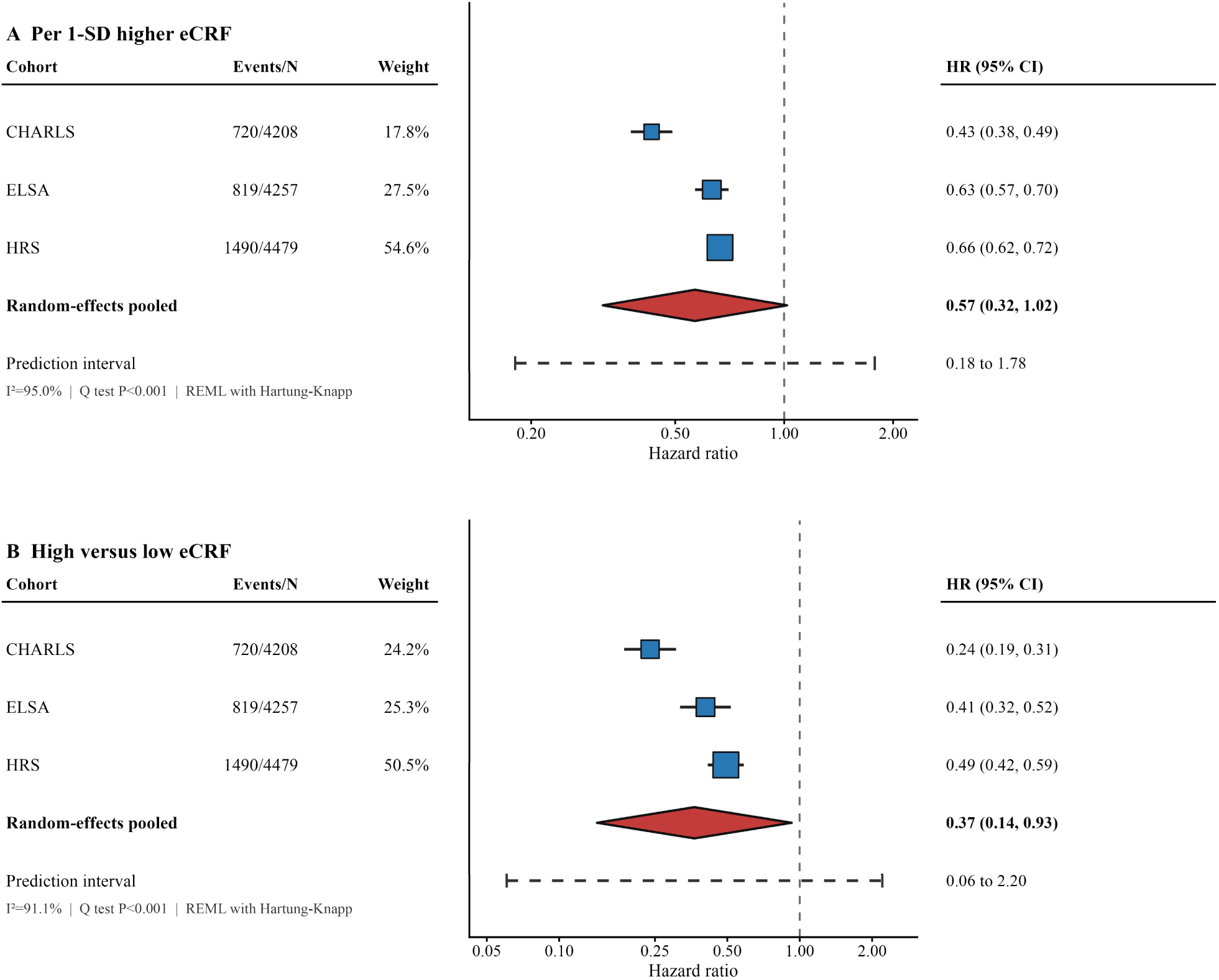
Two-stage random-effects meta-analysis of cohort-specific associations between eCRF and incident CMM. Cohort-specific fully adjusted estimates were pooled by restricted maximum likelihood; Hartung-Knapp intervals and prediction intervals are shown to reflect the small number of cohorts and between-cohort heterogeneity. CMM, cardiometabolic multimorbidity; eCRF, estimated cardiorespiratory fitness; HR, hazard ratio.

**Table 4.** Incremental prognostic performance after adding eCRF.

| Cohort | Time, years | Model 3 AUC (95% CI) | Model 3 + eCRF AUC (95% CI) | AUC increment (95% CI) | IDI (95% CI) | Continuous NRI (95% CI) |
| --- | --- | --- | --- | --- | --- | --- |
| CHARLS | 5 | 0.642 (0.610, 0.674) | 0.702 (0.672, 0.732) | 0.060 (0.033, 0.088) | 0.029 (0.021, 0.038) | 0.222 (0.138, 0.277) |
| CHARLS | 10 | Not estimable | Not estimable | Not estimable | Not estimable | Not estimable |
| ELSA | 5 | 0.684 (0.652, 0.716) | 0.714 (0.684, 0.743) | 0.029 (0.010, 0.048) | 0.010 (0.006, 0.015) | 0.104 (0.049, 0.169) |
| ELSA | 10 | 0.706 (0.683, 0.729) | 0.735 (0.714, 0.756) | 0.029 (0.015, 0.043) | 0.022 (0.013, 0.032) | 0.154 (0.104, 0.193) |
| HRS | 5 | 0.650 (0.627, 0.674) | 0.686 (0.664, 0.708) | 0.036 (0.019, 0.052) | 0.015 (0.011, 0.019) | 0.151 (0.097, 0.187) |
| HRS | 10 | 0.674 (0.655, 0.693) | 0.698 (0.679, 0.716) | 0.024 (0.010, 0.037) | 0.024 (0.015, 0.033) | 0.139 (0.091, 0.179) |
| Pooled | 5 | 0.658 (0.642, 0.674) | 0.698 (0.683, 0.713) | 0.040 (0.029, 0.051) | 0.015 (0.012, 0.018) | 0.155 (0.122, 0.194) |
| Pooled | 10 | 0.707 (0.694, 0.720) | 0.736 (0.724, 0.748) | 0.029 (0.020, 0.038) | 0.032 (0.026, 0.038) | 0.204 (0.176, 0.226) |

Panel B. Harrell C-index
| Cohort | Model 3 | Model 3 plus eCRF | Increment after adding eCRF |
| --- | --- | --- | --- |
| CHARLS | 0.619 (0.598, 0.640) | 0.683 (0.663, 0.703) | 0.064 (0.045, 0.083) |
| ELSA | 0.653 (0.634, 0.673) | 0.686 (0.668, 0.704) | 0.033 (0.021, 0.044) |
| HRS | 0.618 (0.603, 0.633) | 0.650 (0.635, 0.664) | 0.031 (0.021, 0.042) |
| Pooled | 0.639 (0.629, 0.649) | 0.676 (0.666, 0.686) | 0.037 (0.030, 0.044) |

Notes: AUC used inverse probability of censoring weighting. IDI and continuous NRI used 500 perturbation replicates. CHARLS 10-year metrics were not estimable.

**Table 5.** Two-stage random-effects meta-analysis of cohort estimates.

| Contrast | Method | Pooled HR (95% CI) | P | I<sup>2</sup>, % | Q test | Prediction interval |
| --- | --- | --- | --- | --- | --- | --- |
| High versus low eCRF | REML | 0.37 (0.24, 0.56) | <0.001 | 91.1 | 22.20 (df=2), P <0.001 | 0.16 to 0.83 |
| High versus low eCRF | REML with Hartung-Knapp | 0.37 (0.14, 0.93) | 0.043 | 91.1 | 22.20 (df=2), P <0.001 | 0.06 to 2.20 |
| Per 1-SD higher eCRF | REML | 0.57 (0.44, 0.74) | <0.001 | 95.0 | 32.00 (df=2), P <0.001 | 0.34 to 0.95 |
| Per 1-SD higher eCRF | REML with Hartung-Knapp | 0.57 (0.32, 1.02) | 0.053 | 95.0 | 32.00 (df=2), P <0.001 | 0.18 to 1.78 |

Notes: Cohort-specific Model 4 log hazard ratios and standard errors were pooled using restricted maximum likelihood. Hartung-Knapp intervals are shown because only three cohorts were available. The prediction interval describes the expected range of effects in a comparable new cohort.

## 4 Discussion

In this harmonised analysis of three nationally representative cohorts of middle-aged and older adults from China, England, and the United States, higher estimated cardiorespiratory fitness was independently associated with a lower risk of incident CMM. The association persisted after adjustment for demographic, socioeconomic, behavioural, functional, and health-related factors, followed a graded pattern across fitness categories, and remained directionally consistent in competing-risk, imputation, common-follow-up, relative-rank, and cohort-omission analyses. eCRF also provided moderate incremental prognostic information beyond sociodemographic and behavioural predictors. At the same time, formal interaction testing and two-stage meta-analysis showed substantial between-cohort heterogeneity, so the pooled association should be interpreted as an overall pattern rather than as evidence of identical effect size across populations.

Cardiorespiratory fitness is a well-established marker of cardiometabolic health and has been recommended as a clinical vital sign [6]. Because directly measured fitness requires exercise testing that is impractical in most settings, non-exercise estimation equations have been developed and validated [12, 25], with estimated fitness showing associations with cardiovascular outcomes comparable to those of directly measured fitness [21, 26]. Previous work established inverse associations between fitness and individual cardiometabolic conditions, including incident type 2 diabetes [9] and cardiometabolic risk [27], as well as cardiovascular and all-cause mortality [8]. More recently, the scope has broadened from single diseases to multimorbidity: in the UK Biobank, higher exercise-tested fitness was associated with a lower risk of progression from health to a first cardiometabolic disease and onward to multimorbidity [11], and with slower accumulation of chronic disease over time [28]. Our findings add evidence from three ageing cohorts that a non-exercise fitness estimate derived from routine variables is associated with the specific outcome of incident cardiometabolic multimorbidity.

Several pathways may contribute to the observed association. Higher fitness is closely linked to greater insulin sensitivity and more favourable glucose and lipid metabolism [30, 31], reduced systemic inflammation and oxidative stress [32], and lower resting heart rate and blood pressure with improved autonomic and cardiac function [33]. Because hypertension, diabetes, heart disease, and stroke arise from overlapping metabolic and vascular processes, higher fitness may relate to lower risk across several cardiometabolic conditions simultaneously and thereby to a reduced tendency for them to cluster. The strength of the association was not identical across cohorts, which may reflect differences in age structure, body composition, disease prevalence, diagnosis, survey timing, and absolute calibration of the eCRF equation. The positive correlations with grip strength-to-weight ratio and the persistence of the association after within-cohort rank transformation reduce concern that absolute calibration alone explains the findings, but they do not constitute formal validation against measured oxygen uptake.

These observations may have public health relevance. Fitness is modifiable, and aerobic, resistance, and high-intensity interval training can improve fitness and cardiometabolic risk factors in middle-aged and older adults [38–40], while a physically active lifestyle is associated with longer life expectancy even among people who already have cardiometabolic multimorbidity [2]. The progression from a first cardiometabolic condition to multimorbidity is also shaped by modifiable behavioural factors [41]. This study has several strengths, including the use of three large nationally representative cohorts, harmonised measures, identical model structures, prospective follow-up, and extensive sensitivity analyses requested during review. Several limitations should also be acknowledged. First, cardiorespiratory fitness was estimated rather than directly measured; the construct-validity analysis supports plausibility but cannot replace direct validation against measured oxygen uptake. Second, conditions defining CMM were based on self-reported physician diagnoses, which may introduce information bias. Third, complete-case analysis may introduce selection bias, although missingness audits and multiple imputation showed similar estimates. Fourth, death timing and CMM ascertainment were harmonised at the survey-wave level rather than exact dates, so competing-risk findings should be interpreted as sensitivity analyses. Fifth, substantial between-cohort heterogeneity was present, and Hartung-Knapp meta-analytic intervals were wide. Finally, residual confounding and reverse causation cannot be excluded, and the observational design precludes causal inference.

## 5 Conclusion

In three nationally representative cohorts from China, England, and the United States, higher estimated cardiorespiratory fitness was independently associated with a lower risk of incident CMM and provided modest incremental prognostic information. The association was robust across multiple sensitivity analyses, but heterogeneity across cohorts indicates that transportability and calibration require further study. Whether improving fitness reduces CMM incidence warrants confirmation in future intervention studies.

## Author Contributions

Huan Feng contributed to conceptualization, methodology, formal analysis, investigation, writing – original draft, and funding acquisition; Tianyue Zhao contributed to methodology, formal analysis, software, visualization, and writing – original draft; Zhengwei Xie contributed to data curation, validation, and writing – review & editing; and Shanshan Li contributed to conceptualization, supervision, project administration, and writing – review & editing. All authors have read and approved the final version of the manuscript.

## Availability of data and materials

The data analysed in this study are publicly available from the three cohort repositories and were obtained following each study’s standard application and access procedures. The China Health and Retirement Longitudinal Study (CHARLS) data are available from the CHARLS official website (http://charls.pku.edu.cn/). The English Longitudinal Study of Ageing (ELSA) data are available through the UK Data Service (https://ukdataservice.ac.uk/), study number 5050. The Health and Retirement Study (HRS) data are available from the HRS website (https://hrsdata.isr.umich.edu/). The harmonised datasets used in this study were obtained from the Gateway to Global Aging Data (https://g2aging.org/). The analysis code is available from the corresponding author upon reasonable request.

## Clinical trial number

Not applicable.

## Ethics approval and consent to participate

This study is a secondary analysis of de-identified, publicly available data and did not require additional ethical approval; each of the three cohorts obtained ethical approval and written informed consent in its original study. The Health and Retirement Study is sponsored by the National Institute on Aging (grant number NIA U01 AG009740) and is conducted by the University of Michigan (IRB Protocol: HUM00061128). Ethical approval for the English Longitudinal Study of Ageing waves used in this study was granted by NHS Research Ethics Committees under the National Research and Ethics Service (NRES): wave 2 and wave 3 received ethical approval from the London Multi-Centre Research Ethics Committee on 12th August 2004 (MREC/04/2/006) and 27th October 2005 (05/MRE02/63); wave 4 received ethical approval from the National Hospital for Neurology and Neurosurgery & Institute of Neurology Joint Research Ethics Committee on 12th October 2007 (07/H0716/48); wave 5 received ethical approval from the Berkshire Research Ethics Committee on 21st December 2009 (09/H0505/124); wave 6 and wave 7 received ethical approval from the NRES Committee South Central–Berkshire on 28th November 2012 (11/SC/0374) and 28th November 2013 (13/SC/0532); and wave 8 received ethical approval from the South Central – Berkshire Research Ethics Committee on 23rd September 2015 (15/SC/0526). The China Health and Retirement Longitudinal Study was approved by the Biomedical Ethics Review Committee of Peking University (IRB00001052-11015), and all participants provided written informed consent.

## Consent for publication

Not applicable.

## Acknowledgments

We thank the China Health and Retirement Longitudinal Study (CHARLS), the English Longitudinal Study of Ageing (ELSA), and the Health and Retirement Study (HRS) teams for providing the data, and all participants and staff for their contributions to these studies.

## List of abbreviations

ADL: Activities of daily living
BMI: Body mass index
CHARLS: China Health and Retirement Longitudinal Study
CI: Confidence interval
CMM: Cardiometabolic multimorbidity
CRF: Cardiorespiratory fitness
eCRF: Estimated cardiorespiratory fitness
ELSA: English Longitudinal Study of Ageing
HR: Hazard ratio
HRS: Health and Retirement Study
IADL: Instrumental activities of daily living
MET: Metabolic equivalent
OR: Odds ratio
SD: Standard deviation.

